# Women in power: Female leadership and public health outcomes during the COVID-19 pandemic

**DOI:** 10.1101/2020.07.13.20152397

**Authors:** Luca Coscieme, Lorenzo Fioramonti, Lars F. Mortensen, Kate E. Pickett, Ida Kubiszewski, Hunter Lovins, Jacqueline Mcglade, Kristín Vala Ragnarsdóttir, Debra Roberts, Robert Costanza, Roberto De Vogli, Richard Wilkinson

## Abstract

Some countries have been more successful than others at dealing with the COVID-19 pandemic. When we explore the different policy approaches adopted as well as the underlying socio-economic factors, we note an interesting set of correlations: countries led by women leaders have fared significantly better than those led by men on a wide range of dimensions concerning the global health crisis. In this paper, we analyze available data for 35 countries, focusing on the following variables: number of deaths per capita due to COVID-19, number of days with reported deaths, peaks in daily deaths, deaths occurred on the first day of lockdown, and excess mortality. Results show that countries governed by female leaders experienced much fewer COVID-19 deaths per capita and were more effective and rapid at flattening the epidemic’s curve, with lower peaks in daily deaths. We argue that there are both contingent and structural reasons that may explain these stark differences. First of all, most women-led governments were more prompt at introducing restrictive measures in the initial phase of the epidemic, prioritizing public health over economic concerns, and more successful at eliciting collaboration from the population. Secondly, most countries led by women are also those with a stronger focus on social equality, human needs and generosity. These societies are more receptive to political agendas that place social and environmental wellbeing at the core of national policymaking.

## 1. Introduction: policy approaches and the global pandemic

The coronavirus pandemic is impacting daily lives, communities, economies, and exacerbating already existing inequalities. Understanding the pandemic in its full complexity is a difficult task that entails separating intertwined environmental, social, and economic dynamics. Since 1980, the global number of human infectious disease outbreaks has risen, as well as the proportion of vector-borne diseases (Smith et al., 2014). The risk of pandemic outbreaks increases with the loss of natural habitat and biodiversity (IPBES, 2019; Min. Schulze 02/04/2020). Climate change is already affecting vector-borne disease transmission, geographic spread and re-emergence, and its impacts are likely to worsen (Rocklöv and Dubrow, 2020; IPCC, 2018). The outcomes of pandemics depend on how risk-prepared societies and economies are, including levels of population health (Wood and Jóhannsson 2020), health systems, and financial markets. All of this calls for a better understanding of what underpins successful prevention and control, and successful policy choices and implementation.

The study of policy responses to COVID-19 can arguably help us understand how to build future-fit societies, particularly thanks to the heterogeneity of outcomes, which may help clarify which actions and which structural factors may be more significant at determining success in dealing with health crises.

The short-term impacts of COVID-19 can be limited by “flattening the curve” (i.e. reducing the spread) of number of cases over time. A higher peak in number of cases implies a higher risk of overloading health care systems. This in turn causes ineffective treatment for individuals suffering from COVID-19 (and other conditions), leading to a higher number of deaths, greater restrictive measures for longer periods, and eventually generating higher impacts in terms of job losses and economic recession (and their health consequences).

To flatten the curve, most countries adopted lockdown measures, recommending that people stay home, work from home whenever possible, and respect physical distancing. The containment measures, together with fiscal and monetary measures, as well as employment and social measures, differed across countries in terms of the timeliness of the implementation, level of stringency, and extent of interventions (e.g. amount and type of financial aid or income support). In general, countries that implemented emergency measures early on were more successful at limiting contagion and required stricter lockdowns only for a shorter period of time.

Responsiveness to COVID-19 implies early testing, tracing and treating (Sheridan, 2020; Normile, 2020), which also depend on resource capacity. However, in medium to high-income countries the decision to take the pandemic ‘seriously’ was mostly due to political considerations regarding whether economic priorities should trump healthcare concerns. In this regard, some commentators have noted how women leaders were less hesitant than men leaders (Fioramonti et al., 2020; Henley and Roy, 2020; Wittenberg-Cox, 2020).

Against this backdrop, we explore differences in COVID-19 outcomes in terms of number of deaths, number of days with reported deaths, peak in daily deaths, deaths at first day of lockdown, and excess mortality in countries governed by women as opposed to countries led by men. Further, we discuss the possible underlying causes of this relationship.

## 2. Methods and Data

### 2.1 Country selection and measures of COVID-19 impacts

Public data on confirmed cases and deaths from COVID-19 is available from the European Centre for Disease Prevention and Control (ECDC) (http://www.ecdc.europa.eu/ last accessed 31 July 2020). Cases and deaths are reported on a daily basis from December 31, 2019. The total number of cases and deaths from COVID-19 in the ECDC dataset are in accordance with the World Health Organization (WHO) COVID-19 Dashboard (https://covid19.who.int/). From the ECDC dataset, we selected countries with 1) continuous data from December 31, 2019 to June 11, 2020, 2) Gross National Income per capita higher than $3,956 (upper-middle income to high income countries), 3) high to very high Human Development Index (HDI) (which includes life expectancy), and 4) a democratic regime (according to the 2019 Democracy Index). Finally, we excluded countries (Thailand and Sri Lanka) without a distinct peak in daily deaths over the study period. These selection criteria ensure good quality of data and robust cross-country comparisons with regards to the impacts of COVID-19, thus excluding that poverty, lack of liberties or state capacity may determine the differences in outcomes. Furthermore, concerns have been raised by good governance advocates that authoritarian governments may not have been transparent with COVID-19 data, and there is no mechanism for the WHO to verify these numbers (Winter, 2020; The Economist 18 February 2020), hence our decision to only select established democracies. We made one exception to this particular selection criterion for China, for its relevant role as the first country with a COVID-19 outbreak. For each of the 35 countries selected we calculated 1) the count of confirmed deaths from COVID-19 and the mortality rate (deaths/total population), 2) the number of days with at least one reported death, 3) and the highest daily number of deaths over population. Further, we calculated the slope of the curve of deaths, as the ratio of the peak in daily deaths and the number of days from first confirmed death to the day of the peak (Fig. 1).

**Figure 1.**
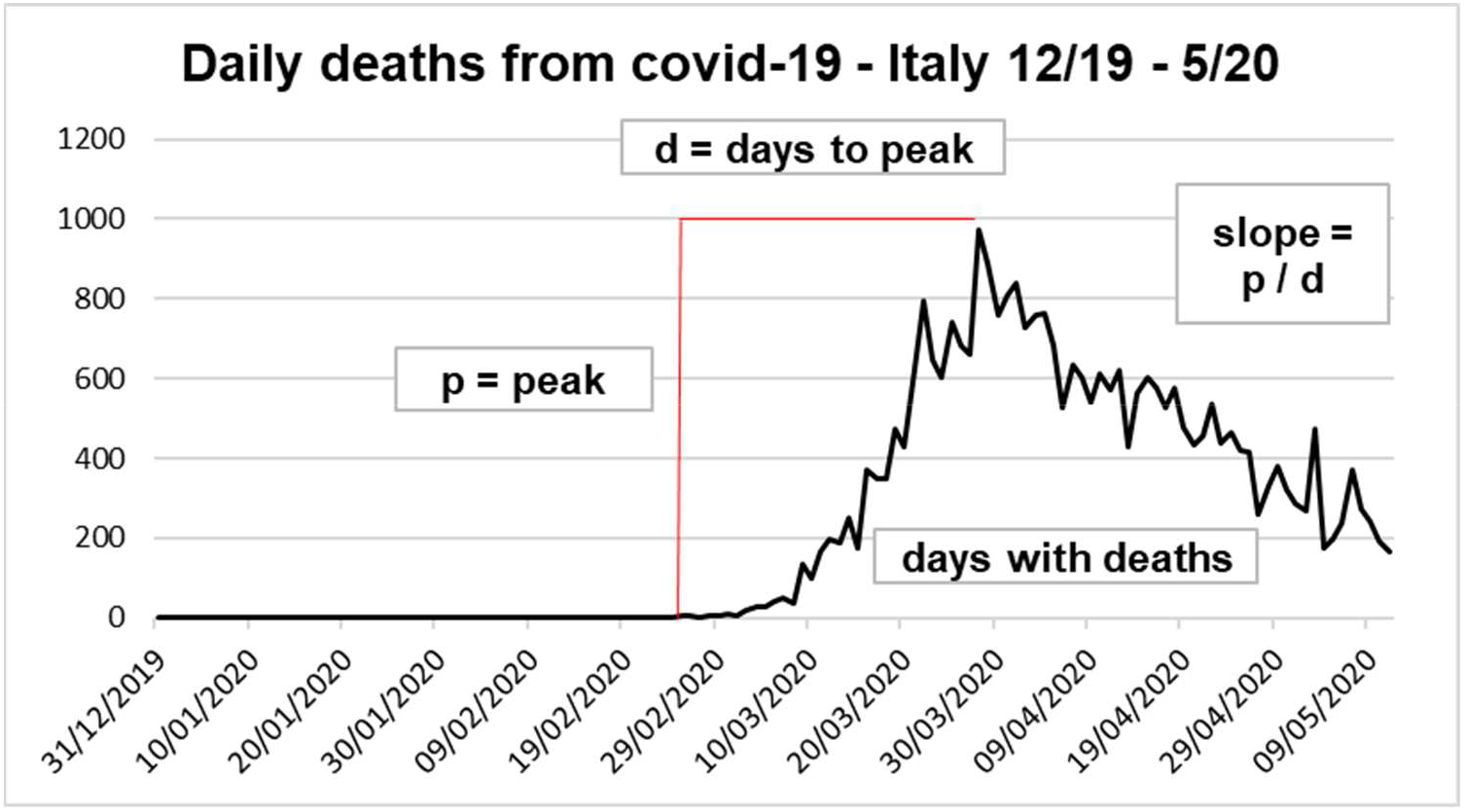
Main measures of impacts of COVID-19 used in this study, using Italy as an example. Days to peak (d) are calculated from first day with deaths. Number of deaths and peak are then divided by population.

As an indicator of the effectiveness and promptness of the policy responses since the outset of the epidemic, we considered the number of deaths in the population at first day of national lockdown. We excluded countries with no lockdown or with only sub-national lockdowns in place to ensure consistency across countries. In order to control for levels of mortality, we analyzed excess mortality from the Financial Times database on “excess mortality during the COVID-19 pandemic” compiled from multiple official sources (the full dataset is available at https://github.com/Financial-Times/coronavirus-excess-mortality-data). From this database, we extracted weekly data of excess mortality and excess of deaths per capita for 18 countries between December 31, 2019 and June 11, 2020.

We grouped countries by the gender of the head of state and government, also considering leaders elected and appointed by a governing committee or parliament where heads of state or government are not directly elected by citizens, excluding women chosen by a hereditary monarch. Of the 35 countries considered, 10 have a woman-led government (Belgium, Denmark, Estonia, Finland, Germany, Greece, Iceland, New Zealand, Norway and Taiwan) while 25 a male-led government (Australia, Austria, Brazil, Canada, China, Croatia, Czechia, Ecuador, France, Ireland, Israel, Italy, Japan, Lithuania, Luxembourg, Mexico, Netherlands, Romania, Russia, South Korea, Spain, Sweden, Switzerland, UK, USA).

### 2.2 Measures of social performance and inequality

Some of the countries currently led by women are also those with the highest global standards in terms of social progress. This is why we have used the Social Progress Index (SPI) 2019 total score, as well as the score of its three main components, namely Basic Human Needs, Foundations of Wellbeing, and Opportunity. Each of these components include four sub-dimensions with three to five indicators each (please refer to https://www.socialprogress.org/ for data and the full list of indicators). In order to explore possible relations between female leadership, impacts of COVID-19, and economic inequality, we have used the Gini coefficient, as well as the income share held by poorest 10% of the population (both from https://data.worldbank.org/). To focus further on gender equality, we have used the Gender Inequality Index (GII) (available from https://hdr.undp.org/), which measures gender inequalities in reproductive health, measured by maternal mortality ratio and adolescent birth rates; empowerment, measured by proportion of parliamentary seats occupied by females and proportion of adult females and males aged 25 years and older with at least some secondary education; and economic status, expressed as labor market participation and measured by labor force participation rate of female and male populations aged 15 years and older. Being an inequality index, higher values of the GII reflect higher gender disparities.

We explored correlations of the GII with the SPI and its sub-dimensions and the global rank in Happiness and Generosity score (from the World Happiness Report 2019; Helliwell et al., 2019) as measures of subjective wellbeing.

## 3. Results

### 3.1 Countries with women leaders are better at reducing negative impacts of COVID-19

Countries with women in position of leadership have suffered six times as few deaths from COVID-19 than countries with governments led by men. When we normalize the data per population, we find that countries led by women had 1.6-times fewer deaths per capita than their male-dominated counterparts (Fig. 2A). Female-led countries reported 1,983 (± 2,724; 95% CI) deaths, while men-led countries 13,276 (± 9,848; 95% CI), by considering average values. The peak in daily deaths was seven times as low in women-led countries (1.5-times lower per capita), where the average highest number of daily COVID-19 deaths was 91 (± 122; 95% CI) across countries, and 643 (± 435; 95% CI) for men-led countries (Fig. 2B). The number of days with confirmed COVID-19 deaths was, on average, 50 (± 23; 95% CI) days in women-led countries and 79 (± 7; 95% CI) in male-led countries (Fig. 2C). Female-led governments managed to flatten the curve more effectively and faster than male-led governments: the slope of the curve of daily deaths from COVID-19 is 4-times less steep in female-led countries (Fig 2D).

**Figure 2.**
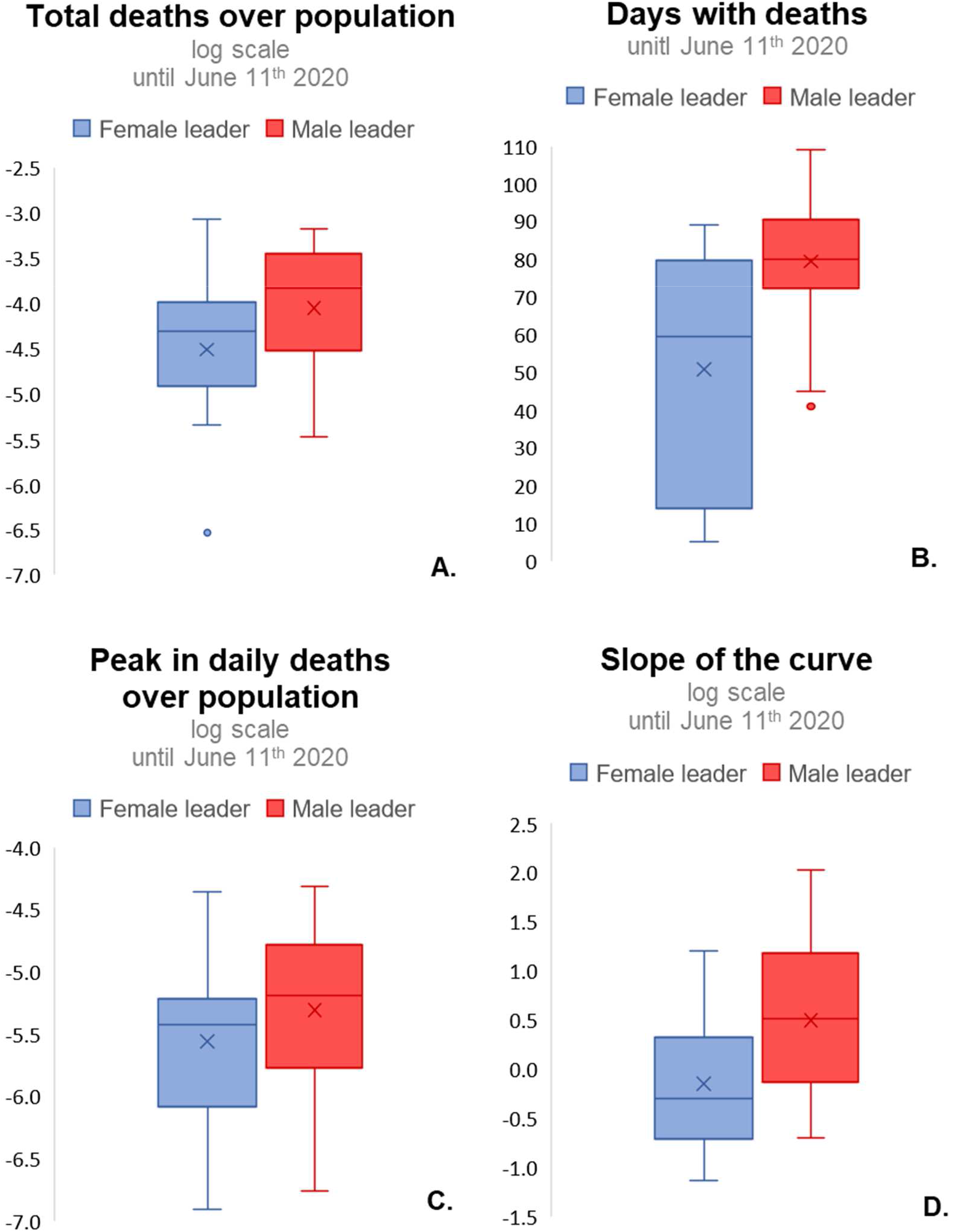
Box-plot of main impacts of COVID-19 in countries with male leaders and countries with female leaders.

As further evidence of different timeliness in implementing emergency response, the average deaths over population at first day of national lockdown was 1.6-times higher in male-led (7.38E-07 ±6.88E-07; 95% CI) than in female-led countries (1.17E-06 ±1.11E-06; 95% CI). Average excess mortality per capita was 4.8 (±13; 95% CI) in female-led countries, and 21 (±24; 95% CI) in men-led countries. This latter result is of particular relevance as excess mortality is acknowledged as the fairest way to compare COVID-19 deaths internationally (Krelle et al., 2020). Furthermore, we found significant positive correlations between deaths over population at first day of lockdown and days with deaths, deaths over population and excess of mortality (Fig. S1)

### 3.2 Impacts of COVID-19 are lower in more equal countries

We found significant positive correlations between economic inequality (higher values of Gini coefficient and lower values of income share held by poorest 10%), and deaths from COVID-19 (total deaths – Fig. 3A, B; and excess mortality – Fig. 3C).

**Figure 3.**
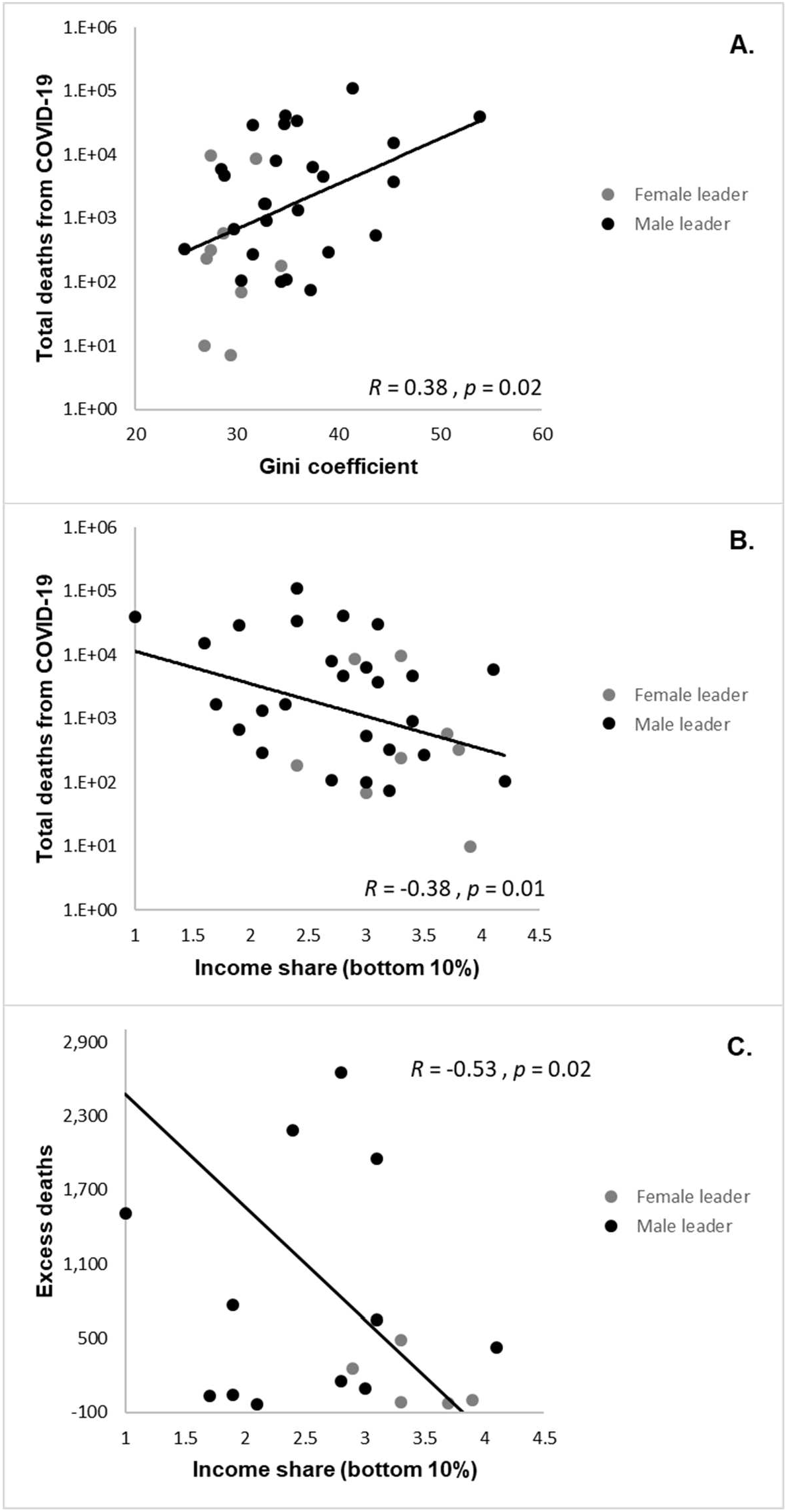
More unequal countries (Gini coefficient) have higher deaths from COVID-19 (A); countries with higher income share held by lowest 10% have lower deaths (B), and lower excess mortality (C), from COVID-19.

Female-led countries have higher scores in all the social progress and equality measures we considered (Fig. 4). The average SPI was 87.87 (± 2.22; 95% CI) in female led-countries, and 81.99 (± 3.16; 95% CI) in men-led countries. Basic Human Needs was 95.46 (± 1.63; 95% CI) in female-led countries, and 91.48 (± 2.22; 95% CI) in men-led countries. Foundations of Wellbeing was 88 (± 2.01; 95% CI) in female-led countries, and 83.6 (± 2.82; 95% CI) in men-led countries. Opportunity was 80.16 (± 3.39; 95% CI) in female-led countries, and 70.89 (± 4.44; 95% CI) in men-led countries. The Gini coefficient was 29.2 (± 1.9; 95% CI) in female-led countries, and 35.7 (± 2.5; 95% CI) in men-led countries. The GII was 0.07 (± 0.02; 95% CI) in female-led countries, and 0.15 (± 0.04; 95% CI) in men-led countries. The average global rank in happiness score was 21 in female-led countries, and 32 in men-led countries, and the rank in generosity was 44 in female-led countries and 59 in men-led countries.

**Figure 4.**
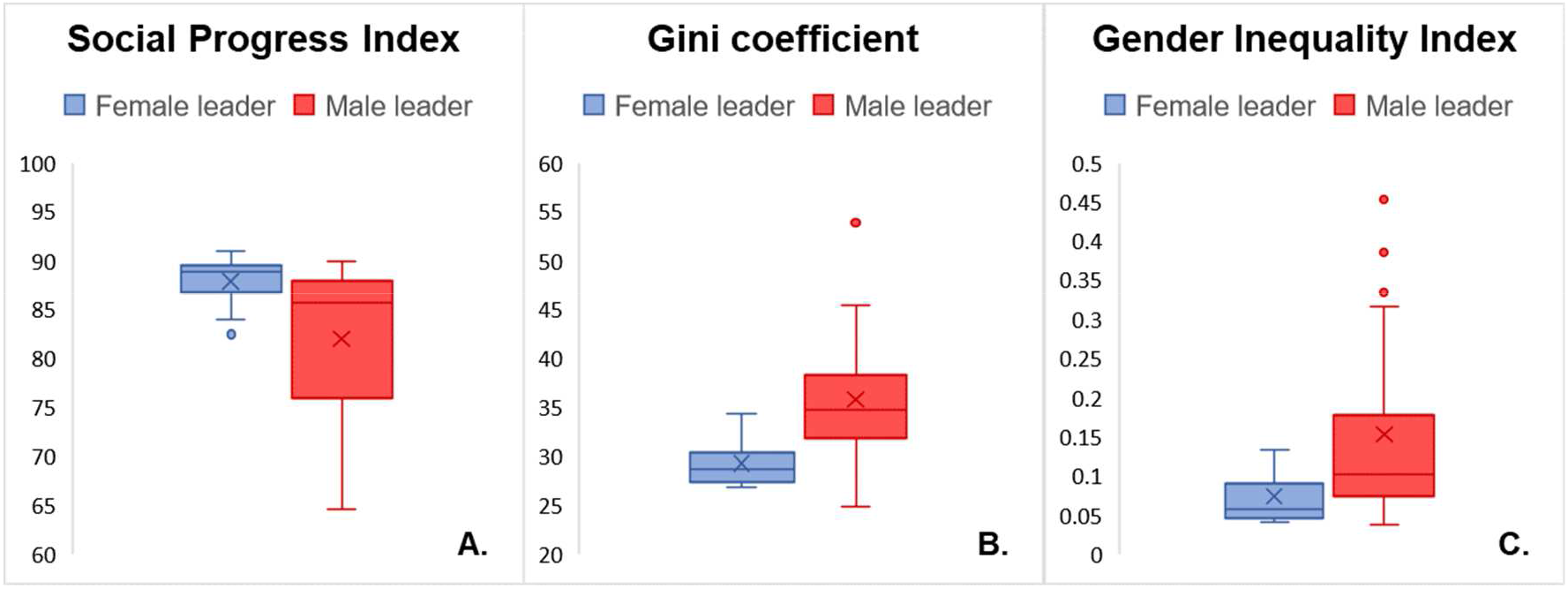
Box-plot of Social Progress Index (A), Gini coefficient (B) and Gender Inequality Index (C) in countries with male leaders and countries with female leaders.

These results point to female leadership as a marker for healthier and more equal societies, where policymaking prioritizes long-term social wellbeing over short-term economic considerations.

Our results with regards to the GII further confirm the relationship between (gender) equality and social well-being. We found that countries with higher female participation and lower gender inequality, besides having higher SPI scores (R = −0.8; p = 2.61E-10), are also happier (R = −0.5; p = 0.004) and more generous (R = −0.5; p = 4.81E-04) (Fig. S2).

As two out of three dimensions of the SPI include aspects related to gender equality (i.e. Foundations of Wellbeing includes “Gender parity in secondary enrollment”, and Opportunity includes “Equality of political power by gender”, https://www.socialprogress.org/) we further explored if more gender equal countries perform better on Basic Human Needs (the one SPI dimension not directly including gender equality indicators). We found that higher scores in the Basic Human Needs dimension of the SPI negatively correlate to the GII (R = −0.8; p = 9.93E-13) (Fig. S3).

To explore if female leadership relates with less negative impacts from COVID-19 even among equal countries with good social performance, we repeated the analysis by considering a sub-set of countries with below-average values of Gini coefficient and GII, and above-average values of SPI. The results confirmed the pattern observed across all 35 countries. In particular, among the 18 countries with below-average Gini coefficient, female-led countries have 26 less days with deaths, 1.12-times fewer deaths over population, and 33.5-times lower excess mortality per capita. Among the 25 countries with below-average GII, female-led countries have 17 less days with deaths, 1.6-times fewer deaths over population, and 6.5-times lower excess mortality per capita. Finally, among the 24 countries with above-average SPI, female-led countries have 29 less days with deaths, 1.8-times fewer deaths over population, and 10.8-times lower excess mortality per capita.

## 4. Discussion

From a policy perspective, the COVID-19 crisis has been characterised by three overarching elements: (1) it has threatened the priority goals of the decision-making unit, namely economic growth; (2) it has compressed the time necessary to develop an appropriate decision; and 3) its eruption has taken the members of the decision-making unit by surprise (Hermann, 1979; de Swielande, 2020).

As COVID-19 deaths began to add up, national leaders were faced with an urgent decision: prioritize economic growth and market openness or shift toward people’s wellbeing. Leaders who opted for the former demonstrated a short-term vision and lack of understanding of the fact that social wellbeing (and a healthy environment) is the *basis* for a healthy economy. Our results show that this is the case for most men leaders, while women leaders did not hesitate to adopt precautionary measures, even when they posed immediate economic costs.

George Lakoff (2010) has argued that conservative and liberal/progressive political views stem ultimately from conceptions of the family and the metaphor of government as parent.

Conservative politics corresponds with a hierarchical “strict father” model of the family. In this model the father’s (and by analogy the political leader’s) authority is absolute and final. The alternative is the “nurturing mother” model corresponding to liberal/progressive politics. The role of the family (and by analogy the government) is to nurture and enable individual and societal progress. “We are all in this together” is an important way of structuring the family and society. When faced with an illness, the strict father might advise working through it while the nurturing mother would advise staying home until you get better.

All modern societies are a mixture of both of these models and they are better adapted to different circumstances. For fighting a war, the hierarchical strict father model works better. But for fighting a pandemic, the nurturing mother model can prove more successful. Countries that lean toward the nurturing mother model of the family and government are more likely to elect progressive female leaders. The fact that countries, such as the United States, supposedly best prepared to fight a pandemic, ended up failing to contain it and suffered more deaths than other nations is evidence of leaders’ failure to take appropriate decisions at the right time. Not taking the COVID-19 crisis seriously led to slow responses and higher social and economic impacts.

In the United States (US), President Donald Trump took much longer than most world leaders to acknowledge the coronavirus crisis (de Swielande, 2020), wasting precious time in managing the crisis and ignoring recommendations from public health experts. Similarly, the UK government overlooked experts calls for early lockdown and the need for widespread and repeated testing (Peto et al., 2020). In Brazil, President Jair Bolsonaro repeatedly called for states to end quarantine measures and fired his health minister Mr. Mandetta, who defended stay-at-home orders (Londoño, 2020; Leonhardt and Leatherby, 2020).

On February 28, 2020, Trump tweeted about COVID-19: “*like a miracle, it will disappear*.” On March 9, his tweet stated: “*The Fake News Media and their partner, the Democrat Party, is doing everything within its considerable power to inflame the Coronavirus situation, far beyond what the facts would warrant*”, clearly downplaying the relevance of the crisis and the urgency for acting.

British PM Boris Johnson missed the first five meetings of the key UK committee on the epidemic, allowing on March 10 to 13 over 250,000 people to gather at the Cheltenham Festival, a clear sign of his underestimation of the crisis and its effects at a time where over 700 cases of COVID-19 were already confirmed in the UK. He visited hospitals and admitted shaking hands and “high-fiving” COVID-19 patients, in a blunt disrespect of any social distancing precautions.

On the opposite side of the spectrum, a number of women leaders heeded scientific active and took immediate action to manage the crisis. Taiwan’s Prime Minister Tsai Ing-wen, building on the country’s previous experience with SARS, introduced targeted measures and medical checks early on, while the epidemic was still in its initial phase in the Chinese city of Wuhan (Wang et al., 2020). This massively reduced the risk of an outbreak and therefore made a lockdown unnecessary. Most other East Asian countries with male leaders, including the equally small Singapore, also affected by SARS in 2002/2003, did not take immediate measures and suffered several waves of contagion.

Iceland’s Prime Minister Katrin Jakobsdottir started crowd restrictions of no more than 20 people gatherings on March 16, 2020. Universities and high-schools went into remote teaching, while primary schools and nurseries were kept open. Businesses were mostly run from employee’s homes. As the number of COVID-19 cases started dropping at the beginning of April, 2020, crowd restrictions became progressively less stringent.

New Zealand’s government of Prime Minister Jacinda Ardern was also prompt in implementing restrictive measures early on, resulting in limited contagion and a much shorter lockdown than neighboring countries in the Pacific. On March 14, New Zealand announced the earliest and toughest self-isolation measures of any country. On the same day, the PM Jacinda Ardern declared “*We’re going hard and we’re going early, … we only have 102 cases, but so did Italy once*.*”* One week later New Zealand was in complete lockdown.

In Scandinavia, the only country that prioritised economic objectives and, as a consequence, did not impose any serious restrictions was Sweden (led by a male prime minister), while all other countries of the region (led my women) took immediate measures. While Norway implemented strict lockdown for almost two months, and Denmark closed upper primary schools (above age 12) from 13 March to 17 May, Sweden opted for a ‘herd immunity’ approach, placing economic priorities ahead of health concerns, keeping primary schools (under age 16) open and only isolating, as much as possible, people over 70 (OCED, 2020). This resulted in the highest COVID-19 mortality rate across Nordic countries by the end of May 2020, with 40.5 deaths per 100,000 population, compared to 9.7 for Denmark and 4.4 for Norway.

The cases above are examples of a more general trend, with female leaders demonstrating more effective management of the pandemic by taking the problem seriously, listening to health experts, and acting quickly. This trend seems to confirm that progressive female leadership is more engaged on issues of health and wellbeing, social equality, sustainability, and innovation, making societies more resilient. Some of these governments have also launched an international alliance to promote, share and further implement wellbeing policies taking the focus off economic growth and putting it on issues that lead to social and ecological wellbeing (https://wellbeingeconomy.org/wego).

In business, there is a tendency for preferential selection of female leaders in times of crisis known as the ‘glass cliff’ effect (Ryan et al., 2011). However, women still represent only 29% of senior leadership in companies (Catalyst 2019). Recognizing the effectiveness of women political leaders in reacting to this coronavirus crisis is one step towards understanding the underlying conditions for effective leadership to emerge.

Implementing policies with short-term economic returns and long-term negative health and social impacts is more common in hierarchical, autocratic societies. These policies often imply pursuing self-interests and attempting to spur dynamics for re-election. There is evidence for women to be more likely to take up positions of political leadership in societies that value equity, solidarity, nurturing, and collaboration, which are usually associated with healthier communities (Wilkinson and Pickett, 2009). Such societal views arose in the 1970s and 80s with the Red Stocking Movement and demand for women being on the lists of political parties and being members of parliaments and local governments (Schneir, 1994).

Women’s status suffers where there is a stronger dominance hierarchy and the “strict father” approach to politics and governance. In more nurturing, sociable (and egalitarian) societies, where position and authority count for less, women’s status tends to be better (Wilkinson and Pickett, 2009). Women’s status is thus a marker for the more egalitarian and sociable societies in which health is less affected by the costs of competition for dominance.

Our results support these points, showing how hierarchical, unequal societies paid higher costs in terms of a broad range of impacts from COVID-19. Our results also show that more equal societies tend to be happier and more generous and tend to better perform better in terms of social progress and environmental quality. Furthermore, even among equal societies, female leaders were more successful than male leaders at dealing with the COVID-19 pandemic.

## 5. Conclusion

We are facing increasing risk of pandemics due to climate change and increasing destruction of ecosystems and biodiversity (IPCC, 2018; IPBES, 2019). While changing our consumption patterns and acting on further drivers of impact is crucial, so it is to build economies and societies that are equal in terms of gender and wealth, with good public health, and are resilient to shocks. The COVID-19 crisis is showing us how political decisions directly affect health and social wellbeing. Women are elected and lead in societies where social and environmental wellbeing is at the core of national policymaking, and this affects a broad range of impacts from COVID-19.

Our results show that female-led countries have consistently less deaths from COVID-19 per capita, a shorter number of days with confirmed deaths, a lower peak in daily deaths per capita, and a lower excess mortality. Female leaders acted quickly, implementing measures of lockdown early on as recommended by national health experts. Our results also show that women are more likely to take up positions of leadership in societies that value equity, nurturing, solidarity, and collaboration, which are usually associated with healthier communities, more resilient to external shocks.

Current data about economic growth forecasts also point out that countries that have taken more determined containment measures will also be rewarded in economic terms: they will suffer much less severe recessions than countries that have hesitated, thus spreading the contagion further afield.

## Data Availability

The data underlying the results presented in the study are available from the European
Centre for Disease Prevention and Control (ECDC) (https://www.ecdc.europa.eu/ last
accessed 31 July 2020) and from the Financial Times database on Excess mortality
during the COVID-19 pandemic, compiled from multiple official sources (the full dataset
is available at https://github.com/Financial-Times/coronavirus-excess-mortality-data).

https://github.com/Financial-Times/coronavirus-excess-mortality-data

## Acknowledgements

The authors are grateful to Katherine Trebeck, Enrico Giovannini and Stewart Wallis for their contributions to this article, and to Amanda Shantz for her contributions on female leadership in business. LC is funded by an IRC/Marie Skłodowska-Curie CAROLINE Postdoctoral Fellowship (IRCCLNE/2017/567).

## References

Catalyst 2019. Quick Take: Women in Management. https://www.catalyst.org/ (August 7, 2019).

Conticini E, Frediani B and Caro D 2020. Can atmospheric pollution be considered a co-factor in extremely high level of SARS-CoV-2 lethality in Northern Italy? Environmental Pollution 261:114465.

de Swielande TS 2020. Trump and Covid-19: The arrogance of ignorance. Commentary paper 68. Université catholique de Louvain.

Fioramonti L, Coscieme L and Mortensen LF 26 May 2020. Women in power: countries with female leaders suffer six times fewer Covid deaths and will recover sooner from recession. Open Democracy. https://www.opendemocracy.net/

Helliwell J, Layard R and Sachs J (Eds.) World Happiness Report 2019; Sustainable Development Solutions Network: New York, NY, USA, 2019.

Henley J and Roy EA 25 April 2020. Are female leaders more successful at managing the coronavirus crisis? The Guardian. https://www.theguardian.com/

Hermann M 1979. Indicators of stress in Policymakers during Foreign Policy Crisis. Political Psychology 1:27–46.

IPBES 2019. Global assessment report on biodiversity and ecosystem services of the Intergovernmental Science-Policy Platform on Biodiversity and Ecosystem Services. Brondizio ES, Settele J, Díaz S, and Ngo HT (Eds). IPBES secretariat, Bonn, Germany.

IPCC 2018. Global warming of 1.5°C. IPCC Special Report on the impacts of global warming of 1.5°C above pre-industrial levels and related global greenhouse gas emission pathways, in the context of strengthening the global response to the threat of climate change, sustainable development, and efforts to eradicate poverty [V. Masson-Delmotte, P. Zhai, H.-O. Pörtner, D. Roberts, J. Skea, P. R. Shukla, A. Pirani, W. Moufouma-Okia, C. Péan, R. Pidcock, S. Connors, J. B. R. Matthews, Y. Chen, X. Zhou, M. I. Gomis, E. Lonnoy, T. Maycock, M. Tignor, T. Waterfield (eds.)]. In Press.

Krelle H, Barclay C and Tallak C 2020. Understanding excess mortality. What is the fairest way to compare COVID-19 deaths internationally? The Health Foundation, 6 May 2020. https://www.health.org.uk/

Lakoff, G., 2010. Moral politics: How liberals and conservatives think. University of Chicago Press.

Leonhardt D and Leatherby L 2 June 2020. Where the virus is growing most: countries with ‘illiberal populist’ leaders. The New York Times. https://nytimes.com/

Londoño E 15 May 2020. Another health minister in Brazil exits amid chaotic coronavirus response. The New York Times. https://nytimes.com/

Min. Schulze (Federal Ministry for the Environment, Nature Conservation and Nuclear Safety, Germany) 2 April 2020. Global Nature conservation can reduce risk of future epidemics. Press release No. 053/20. https://www.bmu.de/

Normile D 17 March 2020. Coronavirus cases have dropped sharply in South Korea. What’s the secret to its success? Science doi:10.1126/science.abb7566

OECD, 2020. Country Policy Tracker. Tackling Coronavirus (COVID-19): Contributing to a global effort. https://www.oecd.org/coronavirus/country-policy-tracker/ (last updated 15 June 2020; last accessed 29 June 2020).

Peto J et al. 2020. Universal weekly testing as the UK COVID-19 lockdown exit strategy. The Lancet 395(10234): 1420–1421 doi:10.1016/S0140-6736(20)30936-3

Rocklöv J and Dubrow R 2020. Climate change: an enduring challenge for vector-borne disease prevention and control. Nature Immunology 21:479–483.

Ryan MK, Haslam SA, Hersby MD and Bongiorno R. 2011. Think crisis–think female: Glass cliffs and contextual variation in the think manager–think male stereotype. Journal of Applied Psychology, 96, 470–484. doi:10.1037/a0022133

Schneir M 1994. Feminism in our time: The essential writings, World War II to Present. Vintage Books, New York.

Sheridan C 23 March 2020. Fast, portable tests come online to curb coronavirus pandemic. Nature Biotechnology 38: 515–518. doi: 10.1038/d41587-020-00010-2

Smith KF et al. 2014. Global rise in human infectious disease outbreaks. J. R. Soc. Interface 11: 201440950. http://dx.doi.org/10.1098/rsif.2014.0950

The Economist, 18 February 2020. Diseases like covid-19 are deadlier in non-democracies. Daily chart. http://economist.com

Wang CJ, Ng CY, and Brook RH 2020. Response to COVID-19 in Taiwan: Big Data Analytics, New Technology, and Proactive Testing. JAMA 323(14): 1341–1342. doi:10.1001/jama.2020.3151

Wilkinson R and Pickett K 2009. The Spirit Level. Penguin Books Ltd.: London, UK. Winter L 2020. Data fog: Why some countries’ coronavirus numbers do not add up. Aljazeera, 17 June 2020. http://aljazeera.com/

Wittenberg-Cox A 13 April 2020. What do countries with the best coronavirus responses have in common? Women leaders. Forbes. https://www.forbes.com/

Wood, T.R. and Jóhannsson, G.F. (2020), Metabolic health and lifestyle medicine should be a cornerstone of future pandemic preparedness. Lifestyle Med.. doi:10.1002/lim2.2

Wu X et al. 2020. Exposure to air pollution and COVID-19 mortality in the United States: A nationwide cross-sectional study. medRxiv https://doi.org/10.1101/2020.04.05.20054502

